# Feeling Too Much or Too Little: Alexithymia Moderates How Self-Reported Physiological Hyperarousal Shapes Risky Decisions

**DOI:** 10.64898/2026.11.02.26367105

**Authors:** Francisco Molins, Blanca Medina-Gascón, Patricia Mesa-Gresa

**Affiliations:** Department of Health Pyschology, Faculty of Health Sciences, Universidad de Alicante, Alicante, Spain; Department of Psychobiology, Faculty of Psychology and Speech Therapy, Universitat de València, Valencia, Spain

**Keywords:** Self-reported physiological hyperarousal, Embodied Cognition, Alexithymia, Risky decision-making, Interoception

## Abstract

**Background:** Embodied cognition frameworks propose that physiological states shape decision-making, yet the contribution of sustained threat-related hyperarousal and individual differences in interoceptive integration remains poorly understood. This study examined whether self-reported physiological hyperarousal predicted Prospect Theory decision parameters: loss aversion (λ), value sensitivity (ρ), and choice consistency (τ); and whether alexithymia moderated these associations.

**Methods:** Sixty-eight healthy young adults (*M* age = 18.94, *SD* = 1.46; 85% women) completed the DASS-21 anxiety subscale as a measure of hyperarousal, the TAS-20 as a measure of alexithymia, and a 64-trial Mixed Gambles Task; individual Prospect Theory parameters were estimated via hierarchical Bayesian analysis. Moderation analyses were conducted using Hayes’ PROCESS macro with Johnson-Neyman probing.

**Results:** Self-reported hyperarousal negatively predicted choice consistency and value sensitivity, but not loss aversion. Alexithymia moderated both effects through distinct conditional patterns: the self-reported hyperarousal–choice consistency association was significant only below a TAS-20 threshold of 51.05 (67.65% of the sample), reflecting monotonic buffering; the self-reported hyperarousal–value sensitivity association was significant within a bounded TAS-20 range (45.31–66.08), suggesting that intermediate alexithymia constitutes a zone of maximal vulnerability, though this model only reached trend significance and should be treated as exploratory. No effects emerged for loss aversion.

**Conclusion:** These findings indicate that sustained hyperarousal could selectively degrade evaluative precision and decision consistency, while leaving loss aversion intact. Alexithymia may operate as a stage-specific, conditional moderator of the body–decision relationship.

## Introduction

Contemporary accounts of embodied cognition and constructivist models of emotion converge on a central claim: cognition cannot be understood independently of the body (Barrett, 2017; Barsalou, 2008; Kiverstein & Miller, 2015; Lindquist et al., 2012). Perception, judgment, and decision-making emerge in close interaction with bodily states, broadly construed to include not only peripheral visceral changes but also the functional state of the brain itself, such as the activation of specific neural networks, metabolic availability, and neuroendocrine regulation (Craig, 2002; Damasio, 1994). From this perspective, cognition, emotion, and physiology are not separable domains, but components of a dynamic system in which fluctuations in bodily state can meaningfully shape information processing and behavior (Barrett, 2017).

Among such bodily states, threat-related hyperarousal is especially relevant. Broadly defined, hyperarousal refers to a state of heightened physiological activation, manifested in symptoms such as accelerated heart rate, agitated breathing, and muscular tension (Chu et al., 2024; Joiner et al., 1996; Ko et al., 2024); and characterized by increased sympathetic nervous system (SNS) activity and neurobiological changes consistent with a stress response, including elevated catecholamines and cortisol, enhanced salience-network recruitment, and a relative reduction in prefrontal control (Arnsten, 2009; Chu et al., 2024; Hermans et al., 2011, 2014). Functionally, this pattern reflects a condition of urgency in which homeostasis is compromised or threatened, and rapid resource mobilization becomes necessary (Clark & Watson, 1991; Joiner et al., 1996), potentially biasing the evaluation of risks, losses, and rewards.

One of the domains that has contributed most to understanding how bodily states shape choices is decision-making under risk. This framework allows the estimation of separable decision parameters, each capturing a distinct psychological dimension of how individuals evaluate and respond to risky options (Sokol-Hessner et al., 2009). Research has mainly focused on three core parameters: loss aversion (λ), the tendency to weight potential losses more heavily than equivalent gains; risk aversion or value sensitivity (ρ), how much an individual differentiates between small and large outcomes; and choice consistency (τ), how reliably an individual’s decisions reflect their own preferences rather than random responding (Sokol-Hessner et al., 2009). Existing findings suggest that threat-related hyperarousal can modulate these parameters in dissociable ways.

Loss aversion has been linked to norepinephrine availability in the brain (Takahashi et al., 2013), and to hyperarousal indexed through electrodermal (Sokol-Hessner et al., 2009) and heart rate activity (Molins et al., 2021). At the same time, hyperarousal induced by an acute laboratory stressor has been associated with reduced risk aversion (Pabst et al., 2013), and experimental reductions in arousal, achieved either through emotion regulation (Sokol-Hessner et al., 2013) or beta-adrenergic blockade with propranolol (Sokol-Hessner, Lackovic, et al., 2015), have been shown to selectively reduce loss aversion while leaving value sensitivity and choice consistency unaffected. In contrast, glucocorticoid activation has been linked to decreased loss aversion (Chumbley et al., 2014; Margittai et al., 2018), greater risk propensity (Pabst et al., 2013), and higher choice consistency (Molins et al., 2023). Together, these findings suggest that the body–cognition link may depend not merely on the presence of hyperarousal, but on its specific configuration and duration.

In this line, an important but underexplored distinction concerns the temporal profile of hyperarousal. Most research has focused on acute, stimulus-bound forms of activation, yet much less is known about the effects of a more diffuse, sustained physiological hyperactivation closer to the elevated bodily tone observed in anxious states. This distinction matters: brief threat-related arousal increases may support momentary mobilization mediated by the central amygdala, a key node in loss aversion responses (Molins & Serrano, 2019; Sokol-Hessner & Rutledge, 2019). By contrast, sustained threat-related hyperarousal appears to engage preferentially the bed nucleus of the stria terminalis and the anterior insula (Alvarez et al., 2011; Davis et al., 2009) and corresponds to elevated tonic locus coeruleus–norepinephrine activity, which can degrade focused, task-driven processing (Aston-Jones & Cohen, 2005). This shift from amygdala-mediated phasic responses toward tonically elevated, diffuse arousal suggests that sustained hyperarousal may impair the precision of value computations and the consistency of choice patterns, potentially affecting value sensitivity and choice consistency more than loss aversion per se.

Furthermore, the influence of hyperarousal on cognition may depend on how bodily signals are perceived and interpreted. Interoceptive sensitivity renders physiological signals more salient and may amplify their impact on judgment, as demonstrated by studies linking heartbeat detection accuracy to greater loss aversion (Sokol-Hessner, Hartley, et al., 2015) or related phenomena, as framing effect (Manzoor et al., 2021). However, detecting bodily signals does not imply accurate interpretation. Alexithymia, a trait characterized by difficulty distinguishing feelings and an externally oriented cognitive style (Nyklícek et al., 2004; Taylor et al., 1997), may reflect a distinct, downstream process. While some approaches suggest that alexithymia could represent a general deficit of interoception (Brewer et al., 2016; Herbert et al., 2011), two-stage models suggest that interoception and alexithymia capture partially independent stages, detection of the bodily signal and its integration into emotional and cognitive experience, respectively (Manzoor et al., 2021; Puigcerver et al., 2024). From this perspective, alexithymia could attenuate the impact of hyperarousal on decision-making, not because the body fails to respond, or because those signals are not consciously processed, but because those they are less effectively incorporated into ongoing cognition, favoring “colder” choices that are less influenced by somatic markers (Bechara & Damasio, 2005; Bibby & Ferguson, 2011).

Consistent with this view, higher alexithymia has been associated with reduced loss aversion (Bibby & Ferguson, 2011), and alexithymia buffered the relationship between COVID-19-related distress and increased loss aversion (Molins & Serrano, 2023). However, prior findings have been limited to loss aversion. Whether alexithymia also moderates the effects of hyperarousal on broader dimensions of risky decision-making, such as value sensitivity and choice consistency, remains unknown.

The present study addressed these gaps. We examined whether sustained threat-related hyperarousal was associated with the variability in loss aversion (λ), value sensitivity (ρ), and choice consistency (τ) estimated from the Mixed Gambles Task (Sokol-Hessner et al., 2009; Tom et al., 2007), and whether alexithymia moderated these associations. Based on the theoretical framework outlined above, we expected sustained hyperarousal to predict reduced value sensitivity and reduced choice consistency, reflecting degraded evaluative precision and increased decision noise (Aston-Jones & Cohen, 2005). Regarding loss aversion, although the distinction between acute and sustained arousal introduces some theoretical ambiguity, the only prior studies examining real and persistent stressors found increases on loss aversion (Belhouk-Herrero et al., 2023; Molins & Serrano, 2023), leading us to predict the same direction here. Moreover, we hypothesized that all associations would be attenuated in individuals with higher alexithymia.

## Method

### Participants

Sample size was determined a priori using G*Power 3.1. Based on effect sizes reported in prior studies examining the relationship between anxiety and decision-making parameters under risk (e.g., Pabst et al., 2013), a small-to-medium effect size was anticipated (f² = 0.15–0.20) for the primary moderation analyses (linear multiple regression, k = 3 predictors, α = .05). This range corresponded to target sample sizes of 60–77 participants to achieve 80% statistical power. A target of 70 participants was therefore established.

Seventy healthy young adults were recruited through convenience sampling from the Faculty of Psychology and Speech Therapy at the Universitat de València, Spain. Two participants were excluded from analyses due to incomplete data resulting from technical errors during administration of the computerized task, yielding a final sample of 68 (58 women, 10 men; *M* age = 18.94 years, *SD* = 1.46, range: 18–23), which remained within the a priori target range and provided approximately 74–80% power across the anticipated effect size range. The sample was predominantly of Spanish origin (85.51%), with a medium-to-high socioeconomic level (*M* = 3.10, *SD* = 0.52). Exclusion criteria included any current neurological or psychiatric diagnosis, habitual drug use, and substance consumption (alcohol or other drugs) within the 24 hours prior to the experimental session. All participants provided written informed consent. The study was approved by the Ethics Committee of the Universitat de València (Reference: 2023-PSILOG-2558999), in accordance with the Declaration of Helsinki.

### Measures

#### Hyperarousal

The anxiety subscale of the Depression Anxiety Stress Scales–21 (DASS-21; Lovibond & Lovibond, 1995; Spanish version by Daza et al., 2002) was used. This subscale comprises 7 items rated on a 4-point Likert scale (0–3). The DASS-21 anxiety subscale specifically targets autonomic arousal and fearfulness, the self-reported physiological hyperarousal component of the tripartite model (Brown et al., 1997; Clark & Watson, 1991), through items indexing symptoms such as trembling, breathing difficulties, and heart pounding. The scale demonstrates adequate psychometric properties in non-clinical Spanish-speaking populations (α = .90; Bados et al., 2005).

#### Alexithymia

The Toronto Alexithymia Scale-20 (TAS-20; Bagby et al., 1994; Spanish version by Martínez-Sánchez, 2014) was used to assess alexithymia. The TAS-20 contains 20 items rated on a 5-point Likert scale (1–5), yielding a total score where higher values indicate greater alexithymia. Established cutoffs classify individuals as non-clinical alexithymic (≤51), possibly clinical alexithymic (52–60), or clinical alexithymic (≥61). The instrument demonstrates adequate reliability and validity in Spanish populations (α = .78; Martínez-Sánchez, 1996).

#### Decision-making under risk

A reduced version of the Mixed Gambles Task (MGT; Chandrasekhar Pammi et al., 2015) was administered. In each of 64 trials, participants were presented with a gamble offering a potential gain and a potential loss and decided whether to accept or reject it (see Figure 1). Following gamble ranges used by Chandrasekhar Pammi et al. (2015), as well as by Tom et al. (2007) in the original task, gains could range from €100 to €380 in €40 increments, and losses from €50 to €190 in €20 increments. In each trial there was a 50% chance of gaining and 50% chance of losing. Individual Prospect Theory parameters were estimated using the computational model described by Sokol-Hessner et al. (2009).

**Figure 1.**
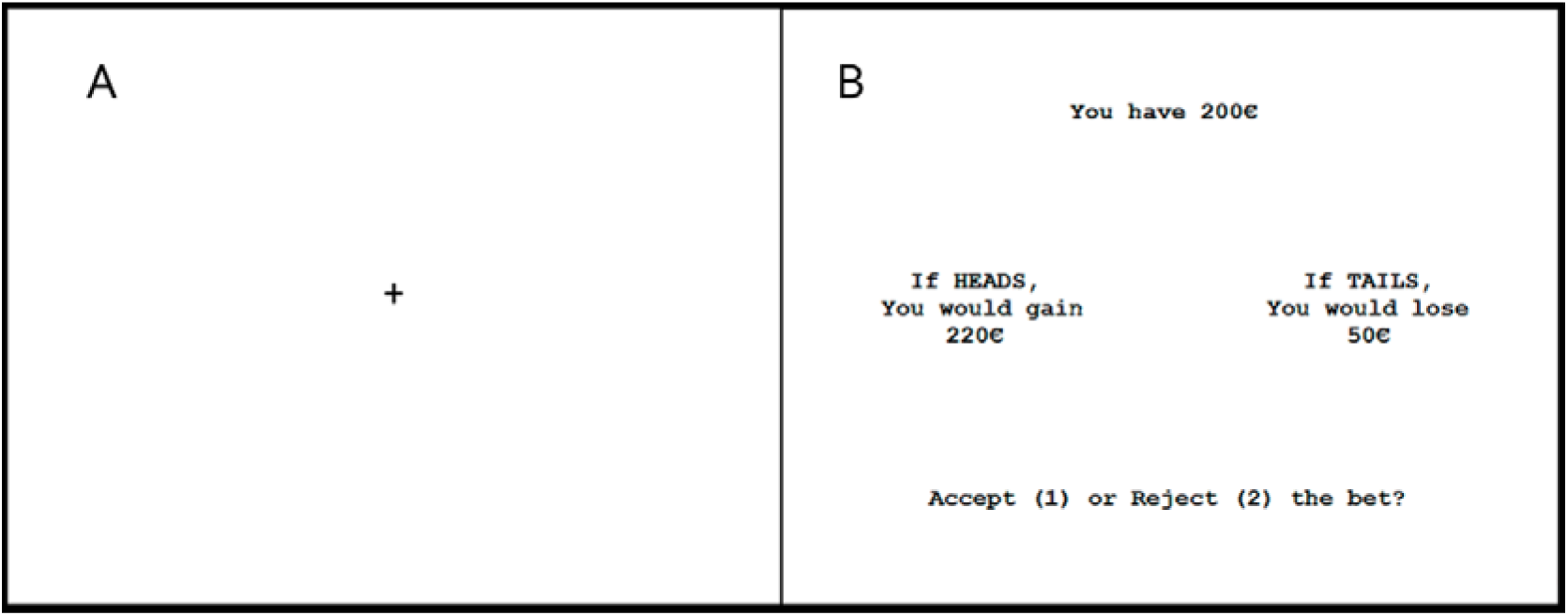
A representative Mixed Gambles Task trial. Each of the 64 trials in the task consists of (A) 5 seconds of fixation point and (B) a bet. The bet offers a possible gain and a possible loss, both with a probability of 50% (heads or tails). The participant must decide whether to play or reject that bet.

#### Prospect-Theory computational model

The Prospect-Theory model (Sokol-Hessner et al., 2009) follows the classical approach of the Prospect Theory (Kahneman & Tversky, 1979) were a bet would be accepted or rejected as a function of the expected utility that it brings to the individual. Following the original paper of Sokol-Hessner et al. (2009), the utility of accepting a bet (U_Accept_) depends on both the utility of the potential gain, estimated through the equation *u(x^gain^) = x*^ρ^; and the utility of the potential loss, estimated through the equation *u(x^loss^) = -* λ *× (-x)*^ρ^. Finally, the probability of accepting a gamble is estimated through the SoftMax function, P_(Accept)_ = 1/(1 + e^−μ(U(Accept)−U(Reject))^); see Sokol-Hessner et al. (2009) for a detailed math description. Three parameters are derived from this model: choice consistency (τ), reflecting the degree to which choices deterministically follow the individual’s utility function (lower values indicate more stochastic, noise-driven choices); value sensitivity (ρ), reflecting the curvature of the subjective value function (lower values indicate greater risk aversion through diminishing marginal sensitivity); and loss aversion (λ), reflecting the relative weight assigned to losses versus gains (values above 1 indicate loss aversion, it is, that losses had a greater impact than gains).

These parameters were estimated for each participant through Hierarchical Bayesian Analyses (HBA; see Ahn et al., 2008 for more details), performed with the hBayesDM package (Ahn et al., 2017) for the R software. The hBayesDM uses Stan 2.1.1 (Stan Development Team, 2017) with the Hamiltonian Monte Carlo (HMC) algorithm as MCMC for sampling the posterior distributions. Following Alacreu-Crespo et al. (2020), we drew 40.000 samples, after burn-in of 23.333 samples, in three different chains (in sum, a total of 120.000 samples and 70.000 burn-in). The Gelman-Rubin test (Gelman & Rubin, 1992) was used to study if the chains converged () to the target distribution. values were 1, which means that convergence was achieved. In addition, to confirm this convergence, the MCMC chains were visually inspected.

### Procedure

Participants individually attended a laboratory session at the Faculty of Psychology and Speech Therapy. Upon arrival, they signed the informed consent form and completed a screening questionnaire verifying inclusion criteria and current state. They then completed the DASS-21 and TAS-20 self-report questionnaires, followed by the computerized MGT. This study was embedded within a broader research protocol collecting additional variables not analyzed here.

### Data Analysis

Outliers were identified using the 2.5 standard deviation method. Assumptions were evaluated using a combination of numerical and graphical procedures. Distributional properties were examined through normality tests and visual inspection of Q–Q plots, while bivariate scatterplots were inspected to assess the plausibility of linear relationships and detect potential extreme cases. For moderation analyses, residual diagnostics were also examined, given that the normality assumption in linear models pertains to the residuals rather than to the raw observed variables. Overall, no severe departures from linearity or other major assumption violations were observed, and regression residuals were consistent with approximate normality.

Descriptive statistics were computed for all study variables. Gender differences were examined using independent-samples *t*-tests. Bivariate correlations were calculated to examine associations between hyperarousal, alexithymia, and the three MGT parameters. To test the moderation hypotheses, three separate regression models were estimated, each predicting one MGT parameter (τ, ρ, or λ) from hyperarousal, alexithymia, and their interaction. We used Hayes’ PROCESS macro for SPSS (Model 1), estimated through ordinary least squares regression with 5,000 bootstrap samples and 95% confidence intervals. The Johnson-Neyman technique (Hayes, 2017; Preacher et al., 2006) was then employed to identify the precise values of the moderator (alexithymia) at which the predictor (hyperarousal) transitions between significant and non-significant effects on the outcome. This approach avoids the arbitrary selection of conditional values inherent in simple slopes analysis. Variables were entered in their original observed metric and were not standardized prior to analysis; therefore, the Johnson–Neyman threshold is expressed in the raw score units of the alexithymia measure. The α significance level was set at .05. All analyses were performed with IBM SPSS Statistics 29.

## Results

### Descriptive Statistics and Gender Differences

Table 1 presents descriptive statistics for all study variables. Mean alexithymia scores indicated that most participants fell below the clinical cutoff for alexithymia (≤51), though sufficient variability existed across the full range. Mean loss aversion values were close to 1, indicating approximate loss neutrality. No significant gender differences were found for any variable (all *p*s > .05), and thus subsequent analyses were conducted on the full sample without gender stratification.

**Table 1.** Descriptive Statistics for Study Variables.

| Variable | Total | Women (*n* = 58) | Men (*n* = 10) | t | p |
|---|---|---|---|---|---|
| Hyperarousal (DASS-21) | 11.78 (3.79) | 11.83 (3.80) | 11.50 (3.89) | 0.25 | .809 |
| Alexithymia (TAS-20) | 47.18 (11.48) | 47.78 (10.74) | 43.70 (15.32) | 0.81 | .437 |
| Choice consistency (τ) | 0.08 (0.02) | 0.07 (0.02) | 0.08 (0.02) | -0.75 | .470 |
| Value sensitivity (ρ) | 2.57 (0.65) | 2.56 (0.64) | 2.63 (0.79) | -0.27 | .789 |
| Loss aversion (λ) | 0.92 (0.01) | 0.92 (0.01) | 0.92 (0.01) | -0.61 | .555 |

Means (Standard deviations).

### Correlations between variables

Table 2 presents the correlation matrix. Hyperarousal and alexithymia were positively correlated, consistent with established literature showing shared variance between these constructs while retaining substantial independence. Regarding decision-making parameters, hyperarousal was negatively correlated with both choice consistency and value sensitivity, indicating that higher hyperarousal was associated with noisier decisions and greater risk aversion; however, it was not significantly related to loss aversion. Moreover, alexithymia was neither significantly correlated with any of the three decision parameters. Among the decision parameters, choice consistency and value sensitivity showed a strong positive correlation, suggesting that individuals with more deterministic choice patterns also exhibit more linear value functions. Finally, value sensitivity and loss aversion were negatively correlated.

**Table 2.** Bivariate Correlations Among Study Variables.

| Variable | 1 | 2 | 3 | 4 | 5 |
|---|---|---|---|---|---|
| 1. Hyperarousal | — |  |  |  |  |
| 2. Alexithymia | .54** | — |  |  |  |
| 3. Choice consistency (τ) | –.27* | –.19 | — |  |  |
| 4. Value sensitivity (ρ) | –.32** | –.15 | .82*** | — |  |
| 5. Loss aversion (λ) | –.08 | –.10 | –.05 | –.36** | — |

Pearson’s Correlation indexes are reported. \* *p* < .05. \*\* *p* < .01. \*\*\* *p* < .001.

### Moderation Analyses

#### Choice consistency (τ)

A moderation model tested whether alexithymia moderated the association between hyperarousal and choice consistency. The overall model accounted for 11.2% of the variance in τ, *F*(3, 64) = 2.68, *p□*= .05, *R*² = .112. Hyperarousal was a significant main predictor (*B* = –0.0043, *SE* = 0.002, *t* = –1.99, *p□*= .049), indicating that higher hyperarousal was associated with lower choice consistency. Alexithymia was not significant as a main effect (*p□*= .112), but the hyperarousal × alexithymia interaction achieved significance (*B* = 0.001, *SE* = 0.0001, *t* = 1.97, *p□*= .05), suggesting that the effect of hyperarousal on choice consistency varies as a function of alexithymia levels. The Johnson-Neyman analysis confirmed this conditional effect: the negative relationship between hyperarousal and choice consistency was significant only for alexithymia values below 51.05 (95% CI [–0.0024, 0.000]), corresponding to 67.65% of the sample (see Figure 2A). For individuals scoring above this threshold on alexithymia, the association between hyperarousal and choice consistency was no longer significant.

**Figure 2.**
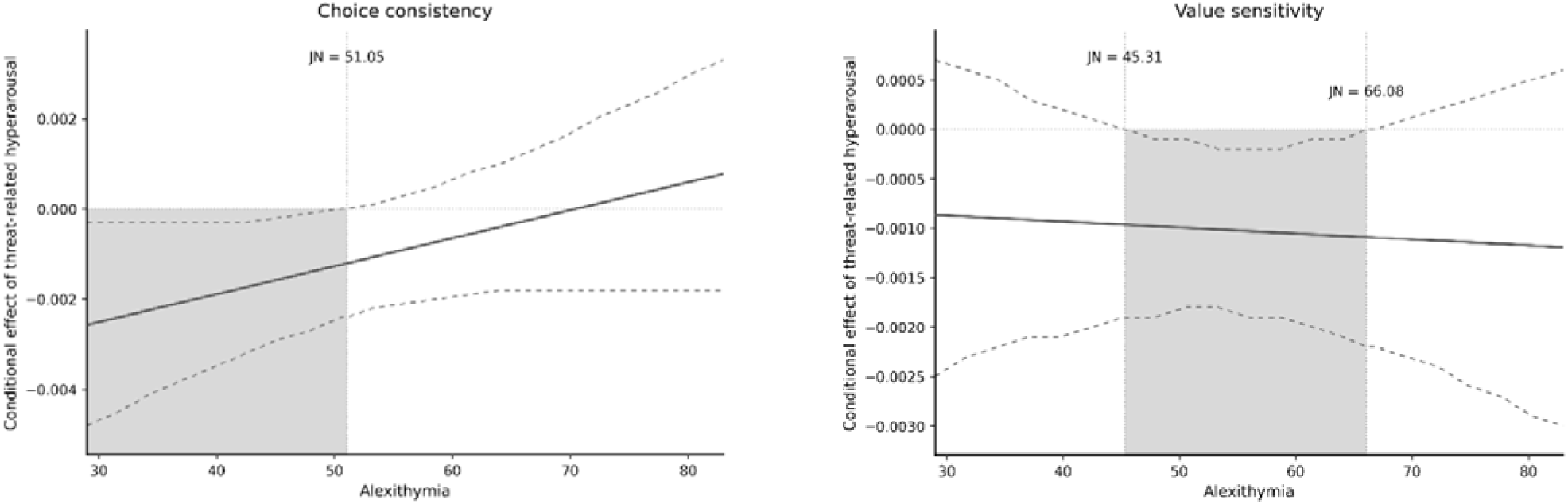
Johnson–Neyman plots of the conditional effect of threat-related hyperarousal on decision-making parameters as a function of alexithymia. Panel A shows the conditional effect of threat-related hyperarousal on choice consistency across levels of alexithymia, whereas Panel B shows the corresponding conditional effect on value sensitivity. The solid line represents the estimated conditional effect of threat-related hyperarousal on each outcome, and the dashed lines indicate the 95% confidence interval. The vertical dotted line(s) mark(s) the Johnson– Neyman (JN) transition point(s), that is, the value(s) of alexithymia at which the confidence interval of the conditional effect intersects zero. The shaded quadrant indicates the region of statistical significance, namely the range of alexithymia scores for which the association between threat-related hyperarousal and the relevant decision-making parameter was statistically significant.

#### Value sensitivity (ρ)

Hyperarousal was also a significant individual predictor of value sensitivity (*B* = –0.007, *SE* = 0.0015, *t* = –2.44, *p□*= .017), suggesting that higher hyperarousal is associated with reduced value sensitivity; that is, greater risk aversion through diminished sensitivity to outcome magnitudes. The hyperarousal × alexithymia interaction was likewise significant (*B* = –0.0001, *SE* = 0.0001, *t* = –2.01, *p□*= .04), and the Johnson-Neyman analysis identified a bounded region of significance for TAS-20 values between 45.31 (95% CI [–0.0019, 0.000]) and 66.08 (95% CI [–0.0022, 0.000]), within which hyperarousal significantly predicted reduced value sensitivity (see Figure 2B). However, the overall regression model only reached a trend of significance, *F*(3, 64) = 2.46, *p□*= .071, *R*² = .103. Therefore, these results should be interpreted cautiously as exploratory, pending replication with larger samples.

#### Loss aversion (λ)

No predictors were significant for loss aversion (all *p*’s > .05). The regression model was not significant, *F*(3, 64) = 0.40, *p□*= .752, *R*² = .019, explaining only 1.9% of the variance. Neither hyperarousal, nor alexithymia, nor their interaction contributed to predicting loss aversion and the Johnson-Neyman analysis yielded no regions of significance. This null finding indicates that the differential weighting of losses relative to gains was not influenced by hyperarousal, alexithymia, or their interaction in this sample.

## Discussion

The present study examined whether sustained, self-reported symptoms of non-specific threat-related hyperarousal was related to risky decision-making parameters, and whether alexithymia moderated this association. Our hypotheses were partially supported: hyperarousal significantly predicted lower choice consistency (τ) and lower value sensitivity (ρ), and alexithymia moderated both effects, albeit through distinct conditional patterns. By contrast, no effects emerged for loss aversion (λ), which remained unrelated to hyperarousal, alexithymia, or their interaction. Taken together, these findings suggest that the relationship between self-reported physiological hyperarousal and decisions could be parameter-specific, dissociable by arousal type (e.g., acute *versus* sustained), and gated by individual differences in the integration of bodily signals into cognitive processing.

First, the negative association between hyperarousal and choice consistency (τ) could be consistent with the theoretical role of tonic locus coeruleus–norepinephrine (LC-NE) activity in degrading decision precision. One possible interpretation is that symptoms associated with sustained hyperarousal may covary with tonic arousal processes in which elevated noradrenergic gain reduces the signal-to-noise ratio of task-relevant processing non-selectively (Aston-Jones & Cohen, 2005). Kane et al. (2017) provided causal evidence for this mechanism, with chemogenetic stimulation of tonic LC activity in rodents producing task impairments, best explained by increased decision noise rather than any systematic directional bias. The present findings are broadly consistent with this pattern, where hyperarousal predicted noisier, less internally consistent choices without shifting the directional valuation of outcomes. This finding also converges with a recent meta-analysis by Pike & Robinson (2022) across 27 studies showing that anxiety, a state in which physiological hyperarousal is a defining feature, is associated with significantly lower inverse-temperature parameters, the computational analogue of choice consistency (τ), across multiple decision paradigms. Moreover, at the cognitive level, Attentional Control Theory (Eysenck et al., 2007) offers a complementary account of how the elevated arousal characteristic of anxious states could impair the inhibition of task-irrelevant thoughts and the flexible shifting of attention across outcome magnitudes, collectively increasing response variability during deliberation.

In this line, the negative association found between hyperarousal and value sensitivity (ρ) extends the picture from pure decision noise to a compression of the subjective value function. Using the same Prospect Theory parameterization employed here, Charpentier et al. (2017) demonstrated that pathological anxiety selectively reduced value sensitivity while leaving loss aversion intact, establishing a behavioral dissociation that our data replicate transdiagnostically in a non-clinical sample. One speculative neuroendocrine interpretation involves processes associated with prolonged glucocorticoid activation. Kandasamy et al. (2014) showed in a controlled pharmacological study that chronically elevated cortisol over 8 days, but not acute administration, reduced value sensitivity and increased risk aversion, a temporal dependency that aligns well with the DASS-21’s sensitivity to persistent physiological tone rather than momentary reactivity. The strong positive correlation between τ and ρ observed in our data is also interpretable in this light. In part, it reflects a well-documented structural interdependency in Prospect Theory models, where choice consistency depends mathematically on value sensitivity (Krefeld-Schwalb et al., 2021). Yet the correlation may also carry substantive meaning: both parameters plausibly reflect a common precision of value representation, such that the neural noise generated by tonic LC-NE activity simultaneously compresses outcome sensitivity and increases choice stochasticity through a shared upstream mechanism.

The moderation analyses refined this picture considerably. Alexithymia moderated the hyperarousal–choice consistency association in a monotonically buffering fashion. The negative effect of hyperarousal on choice consistency was significant only for individuals below a TAS-20 threshold of 51.05, corresponding to approximately two-thirds of the sample. This pattern aligns with the two-stage interoception model (Manzoor et al., 2021), which distinguishes the detection of bodily signals from their downstream integration into cognitive and behavioral processes. In individuals with low alexithymia, arousal signals are not only detected but effectively integrated, enabling them to propagate into choice behavior, consistent with the somatic marker framework (Bechara & Damasio, 2005), and with prior demonstrations that emotional processing ability predicts decision biases (Bibby & Ferguson, 2011; Molins & Serrano, 2023). In higher-alexithymia individuals, the integration stage appears disrupted, severing the body–decision link and rendering physiological arousal computationally inert for that parameter.

However, the moderation pattern for value sensitivity was more complex, and theoretically more revealing. The Johnson-Neyman analysis identified a bounded region of significance (TAS-20 ≈ 45–66), outside which (at both low and high alexithymia), threat-related hyperarousal did not significantly predict value sensitivity. The upper bound seems to replicate the logic above: profound somatic disconnection at high alexithymia prevents arousal from reaching the value computation pathway, consistent with evidence that extreme alexithymia produces selective deficits in emotion-guided decisions without impairing explicit risk evaluation (Zhang et al., 2017). The lower bound is theoretically more interesting. We suggest that individuals with low alexithymia possess sufficient emotion regulation resources to prevent threat-related arousal from distorting their value function, an interpretation potentially supported by experimental demonstrations that reappraisal ability specifically attenuates arousal-linked shifts in risk preferences (Sokol-Hessner et al., 2009, 2013). This positions intermediate alexithymia as a vulnerable zone where arousal is registered but can be neither adequately contextualized nor regulated, producing maximal interference with value computation. Luminet et al. (2021) proposed precisely this intermediate profile, characterized by heightened emotional reactivity without the metacognitive resources to manage it, as theoretically distinct from both intact emotional processing and complete emotional disconnection. The present data provide preliminary behavioral decision-level support for this characterization; however, given that the overall regression model for value sensitivity only reached a trend of significance, these results should be interpreted with caution and treated as exploratory, pending replication with larger samples that afford adequate statistical power to detect moderation effects of this size.

Finally, the null finding for loss aversion (λ) also warrants careful interpretation rather than dismissal. Loss aversion could mostly depend on phasic, stimulus-bound arousal mediated by the central amygdala rather than on the diffuse, sustained hyperarousal captured by the DASS-21. The pharmacological evidence is unambiguous on this point: propranolol selectively reduces λ without affecting ρ or τ (Sokol-Hessner, Lackovic, et al., 2015), and bilateral amygdala damage dramatically attenuates loss aversion while sparing value sensitivity and choice consistency (De Martino et al., 2010). The present null converges with Charpentier et al.’s (2017) finding of no group difference in λ between unmedicated anxious patients and healthy controls, despite robust differences in ρ. Taken together, these findings suggest that prior demonstrations of hyperarousal-linked increases in λ (Molins et al., 2021; Sokol-Hessner et al., 2009) could be better understood as reflecting acute, phasic threat-related hyperarousal rather than sustained hyperarousal indexed here. It is worth noting, however, that the only studies that had previously examined a sustained form of threat exposure (those using COVID-19-related distress), did find increased loss aversion (Belhouk-Herrero et al., 2023; Molins & Serrano, 2023). Rather than contradicting the present null, this divergence may point to a meaningful distinction between psychologically elaborated, emotionally vivid threat, where cognitive appraisal and affective meaning-making actively sustain threat-related processing, and the diffuse self-reported physiological hyperarousal measured here, which may lack the emotional salience necessary to engage the loss aversion mechanism. This interpretation remains speculative, however, and future work directly comparing these forms of arousal within the same paradigm will be needed to adjudicate this empirically.

Despite promising findings, several limitations should be acknowledged. The sample size is adequate for main-effect detection but provides limited power for moderation analyses; the significant interactions must therefore be treated as preliminary pending replication in larger samples, given that interaction effect sizes are typically small relative to main effects (Baranger et al., 2023). Reliance on self-report measures for both hyperarousal and alexithymia introduces shared method variance and precludes direct validation of the hypothesized physiological mechanisms; future studies would benefit from incorporating some objective measures to operationalize hyperarousal independently of subjective report. Finally, the young, predominantly female, non-clinical sample additionally constrains generalizability to populations where these traits exhibit a broader and more consequential range of expression.

In sum, the present study suggests that the relationship between self-reported physiological hyperarousal and risky decision-making is parameter-specific and conditional on the capacity to integrate bodily signals. Higher self-reported hyperarousal may degrade the precision and consistency of value computation, processes sensitive to the diffuse consequences of elevated tonic noradrenergic tone, while leaving loss aversion intact, a parameter whose modulation has previously been linked to phasic amygdala responses, engaged by acute threat. Alexithymia would act not as a uniform buffer but as a stage-specific moderator that showed parameter-specific conditional patterns, attenuating the influence of arousal on choice behavior in a manner that depends on the degree of emotional integration capacity available. These findings underscore the importance of distinguishing between arousal types, decision parameters, and levels of interoceptive processing when examining how bodily states shape decision-making, and call for future work combining objective physiological measures with computational modeling to clarify the pathways through which the anxious body enters the mind.

## Declarations

### Ethics approval

The study was approved by the Ethics Committee of the Universitat de València (Reference: 2023-PSILOG-2558999), in accordance with the Declaration of Helsinki.

### Consent to participate

All participants provided written informed consent.

### Consent for publication

All authors consent for publication.

### Availability of data and materials

The data and analysis code supporting the findings of this study are available from the corresponding author upon reasonable request. This study was not preregistered.

### Code availability

Not applicable.

### Declaration of generative AI and AI-assisted technologies in the writing process

During the preparation of this work the author used Claude in order to improve the English readability. After using this tool, the author reviewed and edited the content as needed and take full responsibility for the content of the publication.

## Data Availability

All data produced in the present study are available upon reasonable request to the authors

## References

Ahn, W. Y., Busemeyer, J. R., Wagenmakers, E. J., & Stout, J. C. (2008). Comparison of decision learning models using the generalization criterion method. Cognitive Science, 32(8), 1376–1402. 10.1080/03640210802352992

Ahn, W. Y., Haines, N., & Zhang, L. (2017). Revealing Neurocomputational Mechanisms of Reinforcement Learning and Decision-Making With the hBayesDM Package. Computational Psychiatry, 1(0), 24. 10.1162/cpsy_a_00002

Alacreu-Crespo, A., Guillaume, S., Sénèque, M., Olié, E., & Courtet, P. (2020). Cognitive modelling to assess decision-making impairments in patients with current depression and with/without suicide history. European Neuropsychopharmacology, 36, 50–59. 10.1016/j.euroneuro.2020.04.006

Alvarez, R. P., Chen, G., Bodurka, J., Kaplan, R., & Grillon, C. (2011). Phasic and sustained fear in humans elicits distinct patterns of brain activity. NeuroImage, 55(1), 389–400. 10.1016/j.neuroimage.2010.11.057

Arnsten, A. F. T. (2009). Stress signalling pathways that impair prefrontal cortex structure and function. Nature Reviews Neuroscience 2009 10:6, 10(6), 410–422. 10.1038/nrn2648

Aston-Jones, G., & Cohen, J. D. (2005). An integrative theory of locus coeruleus-norepinephrine function: Adaptive gain and optimal performance. Annual Review of Neuroscience, 28(Volume 28, 2005), 403–450. 10.1146/annurev.neuro.28.061604.135709

Bados, A., Solanas, A., & Andrés, R. (2005). Psychometric properties of the Spanish version of depression, anxiety and stress scales (DASS). Psicothema.

Bagby, R. M., Parker, J. D. A., & Taylor, G. J. (1994). The twenty-item Toronto Alexithymia Scale—I. Item selection and cross-validation of the factor structure. Journal of Psychosomatic Research, 38(1), 23–32.

Baranger, D. A. A., Finsaas, M. C., Goldstein, B. L., Vize, C. E., Lynam, D. R., & Olino, T. M. (2023). Tutorial: Power Analyses for Interaction Effects in Cross-Sectional Regressions. Advances in Methods and Practices in Psychological Science, 6(3). 10.1177/25152459231187531

Barrett, L. F. (2017). The theory of constructed emotion: an active inference account of interoception and categorization. Social Cognitive and Affective Neuroscience, 12(1), 1–23. 10.1093/scan/nsw154

Barsalou, L. W. (2008). Grounded cognition. Annual Review of Psychology, 59(Volume 59, 2008), 617–645. 10.1146/annurev.psych.59.103006.093639

Bechara, A., & Damasio, A. R. (2005). The somatic marker hypothesis: A neural theory of economic decision. Games and Economic Behavior, 52(2), 336–372. 10.1016/j.geb.2004.06.010

Belhouk-Herrero, M. O., Molins, F., & Serrano, M. Á. (2023). COVID-19 stressor reduces risk taking: the role of trait interoception. Cognitive Processing, 24(3), 353–360. 10.1007/S10339-023-01134-4/FIGURES/2

Bibby, P. A., & Ferguson, E. (2011). The ability to process emotional information predicts loss aversion. Personality and Individual Differences, 51(3), 263–266. 10.1017/s0140525x01224145

Brewer, R., Cook, R., & Bird, G. (2016). Alexithymia: A general deficit of interoception. Royal Society Open Science, 3(10). 10.1098/rsos.150664

Brown, T. A., Chorpita, B. F., Korotitsch, W., & Barlow, D. H. (1997). Psychometric properties of the Depression Anxiety Stress Scales (DASS) in clinical samples. Behaviour Research and Therapy, 35(1), 79–89. 10.1016/S0005-7967(96)00068-X

Chandrasekhar Pammi, V. C., Rajesh, P. P. G., Kesavadas, C., Mary, P. R., Seema, S., Radhakrishnan, A., & Sitaram, R. (2015). Neural loss aversion differences between depression patients and healthy individuals: a functional MRI investigation. Neuroradiology Journal, 28(2), 97–105. 10.1177/1971400915576670

Charpentier, C. J., Aylward, J., Roiser, J. P., & Robinson, O. J. (2017). Enhanced Risk Aversion, But Not Loss Aversion, in Unmedicated Pathological Anxiety. Biological Psychiatry, 81(12), 1014–1022. 10.1016/j.biopsych.2016.12.010

Chu, B., Marwaha, K., Sanvictores, T., Awosika, A. O., & Ayers, D. (2024). Physiology, Stress Reaction. StatPearls. https://www.ncbi.nlm.nih.gov/books/NBK541120/

Chumbley, J. R., Krajbich, I., Engelmann, J. B., Russell, E., Van Uum, S., Koren, G., & Fehr, E. (2014). Endogenous Cortisol Predicts Decreased Loss Aversion in Young Men. Psychological Science, 25(11), 2102–2105. 10.1177/0956797614546555

Clark, L. A., & Watson, D. (1991). Tripartite Model of Anxiety and Depression: Psychometric Evidence and Taxonomic Implications. Journal of Abnormal Psychology, 100(3), 316–336. 10.1037/0021-843X.100.3.316

Craig, A. D. (2002). How do you feel? Interoception: the sense of the physiological condition of the body. Nature Reviews Neuroscience 2002 3:8, 3(8), 655–666. 10.1038/nrn894

Damasio, A. R. (1994). Descartes’ error: Emotion, reason, and the human brain. G.P. Putnam’s Sons.

Davis, M., Walker, D. L., Miles, L., & Grillon, C. (2009). Phasic vs Sustained Fear in Rats and Humans: Role of the Extended Amygdala in Fear vs Anxiety. Neuropsychopharmacology 2010 35:1, 35(1), 105–135. 10.1038/npp.2009.109

Daza, P., Novy, D. M., Stanley, M. A., & Averill, P. (2002). The depression anxiety stress scale-21: Spanish translation and validation with a Hispanic sample. Journal of Psychopathology and Behavioral Assessment, 24, 195–205.

De Martino, B., Camerer, C. F., & Adolphs, R. (2010). Amygdala damage eliminates monetary loss aversion. Proceedings of the National Academy of Sciences of the United States of America, 107(8), 3788–3792. 10.1073/pnas.0910230107

Eysenck, M. W., Derakshan, N., Santos, R., & Calvo, M. G. (2007). Anxiety and cognitive performance: Attentional control theory. Emotion, 7(2), 336–353. 10.1037/1528-3542.7.2.336

Gelman, A., & Rubin, D. (1992). Inference from Iterative Simulation Using Multiple Sequences. Statistical Science, 7(4), 457–511.

Hayes, A. F. (2017). Introduction to mediation, moderation, and conditional process analysis: A regression-based approach. Guilford publications.

Herbert, B. M., Herbert, C., & Pollatos, O. (2011). On the relationship between interoceptive awareness and alexithymia: Is interoceptive awareness related to emotional awareness? Journal of Personality, 79(5), 1149–1175. 10.1111/j.1467-6494.2011.00717.x

Hermans, E. J., Henckens, M. J. A. G., Joëls, M., & Fernández, G. (2014). Dynamic adaptation of large-scale brain networks in response to acute stressors. Trends in Neurosciences, 37(6), 304–314. 10.1016/j.tins.2014.03.006

Hermans, E. J., Van Marle, H. J. F., Ossewaarde, L., Henckens, M. J. A. G., Qin, S., Van Kesteren, M. T. R., Schoots, V. C., Cousijn, H., Rijpkema, M., Oostenveld, R., & Fernández, G. (2011). Stress-related noradrenergic activity prompts large-scale neural network reconfiguration. Science, 334(6059), 1151–1153. 10.1126/science.1209603

Joiner, T. E., Catanzaro, S. J., & Laurent, J. (1996). Tripartite structure of positive and negative affect, depression, and anxiety in child and adolescent psychiatric inpatients. Journal of Abnormal Psychology, 105(3), 401–409. 10.1037/0021-843X.105.3.401

Kahneman, D., & Tversky, A. (1979). Prospect theory: An analysis of decision under risk. Econometrica, 47(2), 263–291. 10.1017/CBO9780511803475.003

Kandasamy, N., Hardy, B., Page, L., Schaffner, M., Graggaber, J., Powlson, A. S., Fletcher, P. C., Gurnell, M., & Coates, J. (2014). Cortisol shifts financial risk preferences Narayanan Kandasamy1. Proceedings of the National Academy of Sciences of the United States of America, 111(9), 3608–3613. 10.1073/pnas.1317908111

Kane, G. A., Vazey, E. M., Wilson, R. C., Shenhav, A., Daw, N. D., Aston-Jones, G., & Cohen, J. D. (2017). Increased locus coeruleus tonic activity causes disengagement from a patch-foraging task. Cognitive, Affective, & Behavioral Neuroscience 2017 17:6, 17(6), 1073–1083. 10.3758/s13415-017-0531-y

Kiverstein, J., & Miller, M. (2015). The embodied brain: Towards a radical embodied cognitive neuroscience. Frontiers in Human Neuroscience, 9(MAY), 1–11. 10.3389/fnhum.2015.00237

Ko, K., Jones, A., Francis, D., Robidoux, S., & McArthur, G. (2024). Physiological correlates of anxiety in childhood and adolescence: A systematic review and meta-analysis. Stress and Health, 40(4), e3388. 10.1002/smi.3388

Krefeld-Schwalb, A., Pachur, T., & Scheibehenne, B. (2021). Structural Parameter Interdependencies in Computational Models of Cognition. Psychological Review, 129(2), 313–339. 10.1037/rev0000285

Lindquist, K. A., Wager, T. D., Kober, H., Bliss-Moreau, E., & Barrett, L. F. (2012). The brain basis of emotion: A meta-analytic review. Behavioral and Brain Sciences, 35(3), 121–143. 10.1017/S0140525X11000446

Lovibond, P. F., & Lovibond, S. H. (1995). The structure of negative emotional states: Comparison of the Depression Anxiety Stress Scales (DASS) with the Beck Depression and Anxiety Inventories. Behaviour Research and Therapy, 33(3), 335–343. 10.1016/0005-7967(94)00075-U

Luminet, O., Nielson, K. A., & Ridout, N. (2021). Cognitive-emotional processing in alexithymia: an integrative review. Cognition and Emotion, 35(3), 449–487. 10.1080/02699931.2021.1908231

Manzoor, N., Molins, F., & Serrano, M. Á. (2021). Interoception moderates the relation between alexithymia and risky-choices in a framing task: A proposal of two-stage model of decision-making. International Journal of Psychophysiology, 162(December 2020), 1–7. 10.1016/j.ijpsycho.2021.01.002

Margittai, Z., Nave, G., Van Wingerden, M., Schnitzler, A., Schwabe, L., & Kalenscher, T. (2018). Combined Effects of Glucocorticoid and Noradrenergic Activity on Loss Aversion. Neuropsychopharmacology: Official Publication of the American College of Neuropsychopharmacology, 43(2), 334–341. 10.1038/npp.2017.75

Martínez-Sánchez, F. (2014). Alexitimia View project Catastrophes, collective behavior and coping View project. (January). https://www.researchgate.net/publication/232447771

Molins, F., Hassen, N. Ben, Paz, M., Costa, R., & Serrano, M. Ã. (2023). Logical decisions after a psychosocial stressor: The late phase of acute stress reduces loss aversion. Physiology and Behavior, 268(November 2022). 10.1016/j.physbeh.2023.114232

Molins, F., & Serrano, M. A. (2019). Bases neurales de la aversión a las pérdidas en contextos económicos: revisión sistemática según las directrices PRISMA. Revista de Neurología, 68(02), 47. 10.33588/rn.6802.2018276

Molins, F., & Serrano, M. Á. (2023). The relationship between the psychological distress derived from COVID-19 and the loss aversion is modulated by the alexithymia trait. Retos, 13(25), 35–46. 10.17163/ret.n25.2023.03

Molins, F., Serrano, M. Á., & Alacreu-Crespo, A. (2021). Early stages of the acute physical stress response increase loss aversion and learning on decision making: A Bayesian approach. Physiology and Behavior, 237(December 2020). 10.1016/j.physbeh.2021.113459

Nyklícek, I., Temoshok, L., & Vingerhoets, A. (2004). Alexithymia: 25 years of theory and research. In Emotional expression and health (pp. 154–170). Routledge.

Pabst, S., Brand, M., & Wolf, O. T. (2013). Stress and decision making: A few minutes make all the difference. Behavioural Brain Research, 250, 39–45. 10.1016/j.bbr.2013.04.046

Pike, A. C., & Robinson, O. J. (2022). Reinforcement Learning in Patients With Mood and Anxiety Disorders vs Control Individuals: A Systematic Review and Meta-analysis. JAMA Psychiatry, 79(4), 313–322. 10.1001/jamapsychiatry.2022.0051

Preacher, K. J., Curran, P. J., & Bauer, D. J. (2006). Computational tools for probing interactions in multiple linear regression, multilevel modeling, and latent curve analysis. Journal of Educational and Behavioral Statistics, 31(4), 437–448. 10.3102/10769986031004437

Puigcerver, M., Molins, F., Molins, M., & Serrano, M. Á. (2024). Interoception moderates the association between alexithymia and anxiety symptoms. Ansiedad y Estrés, 30(1), 35–39.

Sokol-Hessner, P., Camerer, C. F., & Phelps, E. A. (2013). Emotion regulation reduces loss aversion and decreases amygdala responses to losses. Social Cognitive and Affective Neuroscience, 8(3), 341–350. 10.1093/scan/nss002

Sokol-Hessner, P., Hartley, C. A., Hamilton, J. R., & Phelps, E. A. (2015). Interoceptive ability predicts aversion to losses. Cognition and Emotion, 29(4), 695–701. 10.1080/02699931.2014.925426

Sokol-Hessner, P., Hsu, M., Curley, N. G., Delgado, M. R., Camerer, C. F., & Phelps, E. A. (2009). Thinking like a trader selectively reduces individuals’ loss aversion. Proceedings of the National Academy of Sciences of the United States of America, 106(13), 5035–5040. 10.1073/pnas.0806761106

Sokol-Hessner, P., Lackovic, S. F., Tobe, R. H., Camerer, C. F., Leventhal, B. L., & Phelps, E. A. (2015). Determinants of Propranolol’s Selective Effect on Loss Aversion. Psychological Science, 26(7), 1123–1130. 10.1177/0956797615582026

Sokol-Hessner, P., & Rutledge, R. B. (2019). The Psychological and Neural Basis of Loss Aversion. Current Directions in Psychological Science, 28(1), 20–27. 10.1177/0963721418806510

Stan Development Team. (2017). Stan Modeling Language User’s Guide and Reference Manual. Version 2.17.0.

Takahashi, H., Fujie, S., Camerer, C., Arakawa, R., Takano, H., Kodaka, F., Matsui, H., Ideno, T., Okubo, S., Takemura, K., Yamada, M., Eguchi, Y., Murai, T., Okubo, Y., Kato, M., Ito, H., & Suhara, T. (2013). Norepinephrine in the brain is associated with aversion to financial loss. Molecular Psychiatry, 18(1), 3–4. 10.1038/mp.2012.7

Taylor, G. J., Bagby, R. M., Parker, J. D. A., & Grotstein, J. (1997). Disorders of Affect Regulation: Alexithymia in Medical and Psychiatric Illness. Disorders of Affect Regulation. 10.1017/cbo9780511526831

Tom, S. M., Fox, C. R., Trepel, C., & Poldrack, R. A. (2007). The neural basis of loss aversion in decision-making under risk. Science, 315(5811), 515–518. 10.1126/science.1134239

Zhang, L., Wang, X., Zhu, Y., Li, H., Zhu, C., Yu, F., & Wang, K. (2017). Selective impairment of decision making under ambiguity in alexithymia. BMC Psychiatry, 17(1), 1–8. 10.1186/s12888-017-1537-2

